# Do Assisted Living Facilities That Offer a Dementia Care Program Differ from Those That Do Not? A Population-Level Cross-Sectional Study in Ontario, Canada

**DOI:** 10.1101/2021.02.06.21251214

**Authors:** Derek R. Manis, Ahmad Rahim, Jeffrey W. Poss, Iwona A. Bielska, Susan E. Bronskill, Jean-Éric Tarride, Julia Abelson, Andrew P. Costa

## Abstract

**Objective:** To identify the characteristics of licensed assisted living facilities that provide a dementia care program compared to assisted living facilities that do not provide such a program.

**Design, Setting, and Participants:** Population-level cross-sectional study in Ontario, Canada on all licensed assisted living facilities in 2018 (*n* = 738).

**Methods:** Data on facility-level characteristics (e.g., resident and suite capacities, etc.) and the provision of other provincially regulated care services (e.g., pharmacist and medical services, skin and wound care, etc.) attributed to licensed assisted living facilities were examined. Multivariable Poisson regression with robust standard errors was used to model the characteristics of assisted living facilities associated with the provision of a dementia care program.

**Results:** There were 123 (16.7%) assisted living facilities that provided a dementia care program. Nearly half of these facilities had a resident capacity exceeding 140 older adults (44.7%) and more than 115 suites (46.3%). All assisted living facilities that provided a dementia care program also provided nursing services, meals, assistance with bathing and hygiene, and administered medications. After adjusting for facility characteristics and other provincially regulated care services, the prevalence of a dementia care program was nearly three times greater in assisted living facilities that offered assistance with feeding than those that did not (Prevalence Ratio [PR] = 2.91, 95% Confidence Interval [CI] 1.98 to 4.29), and almost twice as great among assisted living facilities that provided medical services than those that did not (PR = 1.78, 95% CI 1.00 to 3.17).

**Conclusions:** A dementia care program was more prevalent in assisted living facilities that housed many older adults, had many suites, and provided at least five of the other 12 regulated care services. These findings deepen the understanding of specialized care for dementia in assisted living facilities.

**BRIEF SUMMARY:** *Question:* Do licensed assisted living facilities that provide a dementia care program differ from those that do not in Ontario, Canada?

*Findings:* Assisted living facilities that provide a dementia care program house many older adults, have many suites, and offer at least five of the other 12 regulated care services.

*Meaning:* Assisted living facilities that provide a dementia care program are larger and provide an array of care services.

*Disclosures:* We have no perceived or actual conflicts, financial or otherwise, to disclose.

*Author Contributions:* DRM and APC conceptualized the study. AR created the dataset, and DRM conducted the statistical analyses. DRM wrote the manuscript, and all authors critically read and contributed to it.

*Data Availability:* The dataset from this study is held securely in coded form at ICES. While data sharing agreements prohibit ICES from making the data set publicly available, access may be granted to those who meet pre-specified criteria for confidential access, available at www.ices.on.ca/DAS. The full data set creation plan and underlying analytic code are available from the corresponding author upon request, understanding that the programs may rely upon coding templates or macros that are unique to ICES.

## INTRODUCTION

Dementia affects more than half of all residents housed in assisted living facilities. ^1–3^ Older adults living with dementia are more likely to experience injuries requiring acute care, be diagnosed with pneumonia, and encounter difficulties with eating. ^4,5^ Care for dementia is expensive and a widely cited reason for older adults requiring placement in a nursing home. ^6–9^ Specialized care for older adults living with dementia, such as a dementia care program, has demonstrated reductions in acute health service use and long-stay transitions to a nursing home. ^8,10^

Assisted living facilities are referred to as retirement homes in the province of Ontario, Canada. Similar to assisted living facilities in the United States that are regulated at the state-level, ^11^ retirement homes are regulated by an independent, not-for-profit regulator (i.e., Retirement Homes Regulatory Authority [RHRA]) in Ontario since 2011. ^12^ The retirement home sector in Ontario has a resident capacity equivalent to that of its nursing home sector (i.e., more than 70,000 older adults), ^12^ which demonstrates that it is a sizeable, important sector providing supportive care to older adults. Unlike nursing homes, residency in a retirement home is exclusively financed through private, out-of-pocket payments by residents and/or their family caregivers. ^12–14^

Much of the literature on dementia care in assisted living facilities addresses health service use among residents living with dementia, managing staff, and state-level regulations for dementia care. ^1,11,15–17^ Studies explicitly investigating the characteristics of retirement homes or assisted living facilities that provide specialized care for dementia (i.e., a dementia care program), and how these characteristics compare to those that do not, are absent. The findings from such studies are important for identifying case mix and examining scope and breadth of care for older adults with complex care needs. A growing proportion of residents of assisted living facilities live with dementia, ^18^ and improving the understanding of dementia care programs in assisted living facilities contributes to informing the sector, community-based dementia care, and national dementia care strategies. The objective of this study is to identify the characteristics of licensed assisted living facilities that provide a dementia care program compared to assisted living facilities that do not provide such a program in Ontario, Canada.

## METHODS

### Study Design and Setting

A population-level cross-sectional study was conducted at ICES in Ontario, Canada. ICES is an independent, non-profit research institute funded by an annual grant from the Ontario Ministry of Health and Long-Term Care. As a prescribed entity under Ontario’s privacy legislation, ICES is authorized to collect and use health care data for the purposes of health system analysis, evaluation, and decision support. Secure access to these data is governed by policies and procedures that are approved by the Information and Privacy Commissioner of Ontario. The REporting of studies Conducted using Observational Routinely-collected health Data (RECORD) statement guideline was followed (Appendix Table 2). ^19^

**Table 1.** Descriptive Characteristics of Licensed Assisted Living Facilities in 2018 (*n* = 738)

|  | **Dementia Care Program** |  | **Standardized Difference** |
| --- | --- | --- | --- |
|  | **Yes** | **No** |  |
| *n* (%) | 123 (16.7) | 615 (83.3) |  |
| **Facility Characteristics, *n* (%)** |  |  |  |
| Urban Location | 114 (92.7) | 508 (82.6) | 0.309 |
| Facility Capacity |  |  | 0.583 |
| 6-49 | 24 (19.5) | 155 (25.2) |  |
| 50-86 | 14 (11.4) | 171 (27.8) |  |
| 87-139 | 30 (24.4) | 156 (25.4) |  |
| 140+ | 55 (44.7) | 133 (21.6) |  |
| Total Suites |  |  | 0.615 |
| 6-41 | 20 (16.3) | 163 (26.5) |  |
| 42-70 | 16 (13.0) | 168 (27.3) |  |
| 71-114 | 30 (24.4) | 156 (25.4) |  |
| 115+ | 57 (46.3) | 128 (20.8) |  |
| Chain Status | 74 (60.2) | 281 (45.7) | 0.293 |
| Residential Home | 8 (6.5) | 71 (11.5) | 0.176 |
| Co-Located with Nursing Home | 19 (15.4) | 112 (18.2) | 0.073 |
| **Care Services, *n* (%)** |  |  |  |
| Assistance with Bathing | 123 (100.0) | 581 (94.5) | 0.342 |
| Assistance with Hygiene | 123 (100.0) | 531 (86.3) | 0.562 |
| Assistance with Ambulation | 117 to 123 (95.1 to 100.0) * | 517 (84.1) | 0.480 |
| Assistance with Feeding | 89 (72.4) | 185 (30.1) | 0.933 |
| Assistance with Dressing | 117 to 123 (95.1 to 100.0) * | 532 (86.5) | 0.507 |
| Continence Care | 117 to 123 (95.1 to 100.0) * | 457 (74.3) | 0.788 |
| Skin and Wound Care | 47 (38.2) | 113 (18.4) | 0.451 |
| Provision of Meals | 123 (100.0) | 609 to 615 (99.0 to 100.0) * | 0.057 |
| Administration of Medications | 123 (100.0) | 609 to 615 (99.0 to 100.0) * | 0.114 |
| Pharmacist Services | 117 to 123 (95.1 to 100.0) * | 535 (87.0) | 0.287 |
| Nursing Services | 123 (100.0) | 574 (93.3) | 0.377 |
| Medical Services | 107 (87.0) | 401 (65.2) | 0.528 |

\* Small cell sizes (i.e., where six or fewer assisted living facilities have, or do not have, a characteristic) are suppressed due to privacy restrictions at ICES

**Table 2.** Associations with the Provision of a Dementia Care Program in Licensed Assisted Living Facilities

|  | Unadjusted PR (95% CI) | Adjusted PR (95% CI) † |
| --- | --- | --- |
| **Facility Characteristics** |  |  |
| Urban | 2.26 (1.23 to 4.52) ** | 1.15 (0.61 to 2.17) |
| Facility Capacity |  |  |
| 6-49 | 1.00 (Reference) | 1.00 (Reference) |
| 50-86 | 0.56 (0.30 to 1.06) | 0.34 (0.18 to 0.66) ** |
| 87-139 | 1.20 (0.73 to 1.98) | 0.43 (0.20 to 0.93) * |
| 140+ | 2.18 (1.41 to 3.37) *** | 0.59 (0.25 to 1.42) |
| Total Suites |  |  |
| 6-41 | 1.00 (Reference) | 1.00 (Reference) |
| 42-70 | 0.80 (0.43 to 1.49) | 1.40 (0.73 to 2.70) |
| 71-114 | 1.48 (0.87 to 2.50) | 2.28 (1.02 to 5.11) * |
| 115+ | 2.82 (1.77 to 4.50) *** | 2.78 (1.09 to 7.07) * |
| Chain Status | 1.63 (1.17 to 2.27) ** | 1.21 (0.88 to 1.67) |
| Residential Home | 0.58 (0.29 to 1.14) | 0.75 (0.35 to 1.61) |
| Co-Located with a Nursing Home | 0.85 (0.54 to 1.33) | 1.21 (0.78 to 1.87) |
| **Care Services** |  |  |
| Assistance with Ambulation | 6.34 (2.05 to 19.57) ** | 0.96 (0.34 to 2.75) |
| Assistance with Feeding | 4.43 (3.07 to 6.39) *** | 2.91 (1.98 to 4.29) *** |
| Assistance with Dressing | 15.67 (2.22 to 110.82) ** | 2.24 (0.26 to 18.96) |
| Continence Care | 33.50 (4.71 to 238.20) *** | 13.51 (1.64 to 111.67) * |
| Skin and Wound Care | 2.23 (1.62 to 3.07) *** | 1.18 (0.85 to 1.63) |
| Pharmacist Services | 2.57 (1.17 to 5.66) * | 0.91 (0.38 to 2.21) |
| Medical Services | 3.03 (1.83 to 5.00) *** | 1.78 (1.00 to 3.17) * |

PR = Prevalence Ratio; CI = Confidence Interval
†Adjusted for all variables in the table
\* $p < 0.05$ ; \*\* $p < 0.01$ ; \*\*\* $p < 0.001$

## Data

A list of all licensed assisted living facilities in Ontario in 2018 was obtained from the public register of the RHRA and imported to ICES (*n* = 757). The postal code of each assisted living facility was linked to Statistics Canada’s Postal Code Conversion file, which is a specialized macro for use with health system administrative datasets containing postal codes. This macro is based on 2016 Census information, flags communities with a population less than 10,000 individuals as rural, and includes related data from Canada Post Corporation. ^20^ Nineteen assisted living facilities (*n* = 19) were removed from the analysis because of missing facility-level and postal code data.

### Exposures

The exposures of interest are facility-level characteristics (i.e., urban location, resident capacity, total suites, chain status, residual home status, and co-location with a nursing home) and the other 12 provincially regulated care services provided in an assisted living facility (Appendix Table 1).

### Outcome

The primary outcome is whether the assisted living facility provided a dementia care program. Dementia care programs in assisted living facilities in Ontario are regulated to include communication strategies, mental stimulation activities, health and wellness monitoring and promotion, and identification of triggers for responsive behaviours. ^21^ These programs must also be supervised by a regulated health care professional (e.g., registered nurse, physician, etc.), align with current evidence and best practices for dementia care, and be evaluated annually. ^21^

### Statistical Analysis

Counts, percentages, and standardized differences were calculated to describe the facility-level and care service characteristics of assisted living facilities that provided, and did not provide, a dementia care program. ^22^ Multivariable Poisson regression with robust standard errors was used to model unadjusted and adjusted estimates with 95% confidence intervals to identify the characteristics of assisted living facilities associated with the provision of a dementia care program. ^23,24^ Tests were two-tailed, and the level of statistical significance was set at α = 0.05. The deviance goodness-of-fit test was calculated to assess whether the Poisson regression model was appropriate. Variance inflation factors were calculated to assess for multicollinearity. ^25^ Data set processing was conducted in SAS Enterprise 9.4 (Cary, NC, USA) and statistical analyses were conducted in Stata MP 16.1 (College Station, TX, USA).

## RESULTS

There were 738 licensed assisted living facilities in Ontario in 2018 (*n* = 738). Of these, 123 provided a dementia care program (16.7% versus 83.3% no dementia care program), and almost all were located in an urban area (92.7% versus 82.6% no dementia care program) (Table 1). Nearly half of these assisted living facilities had a resident capacity of 140 or more (44.7% versus 21.6% no dementia care program) and had more than 115 suites (46.3% versus 20.8% no dementia care program). All assisted living facilities that provided a dementia care program also provided nursing services, meals, assistance with bathing and hygiene, and administered medications (*n* = 123). Many of the standardized differences between assisted living facilities that provided a dementia care program and those that did not exceeded 10%, which indicated that assisted living facilities that provided a dementia care program were systematically different from those that did not. ^22^

Assistance with bathing and hygiene, provision of meals, administration of medication, and nursing services were removed from the adjusted model because of collinearity, and there was no evidence of multicollinearity in the adjusted model (i.e., variance inflation factors equal to or greater than a value of 10). ^25^ The deviance goodness-of-fit statistic was not statistically significant. After adjusting for facility characteristics and regulated care services, the prevalence of a dementia care program was almost three times greater in assisted living facilities with 115 or more suites (Prevalence Ratio [PR] = 2.78, 95% Confidence Interval [CI] 1.09 to 7.07) compared to assisted living facilities with 41 or fewer suites (Table 2). The prevalence of a dementia care program was nearly three times greater in assisted living facilities that provided assistance with feeding (PR = 2.91, 95% CI 1.98 to 4.29), and the prevalence of a dementia care program was almost twice as great in assisted living facilities that provided medical services (PR = 1.78, 95% CI 1.00 to 3.17), compared to assisted living facilities that did not provide these care services.

## DISCUSSION

Assisted living facilities that provided a dementia care program were systematically different from those that did not provide such a program. Specifically, assisted living facilities in Ontario that provided a dementia program had large resident capacities, many suites, and provided, at a minimum, nursing services, meals, assistance with bathing and hygiene, and administered medications. The prevalence of a dementia care program in an assisted living facility was greater in assisted living facilities where assistance with feeding and medical services were also provided.

More than 90% of assisted living facilities that provided a dementia care program were located in urban communities. Consistent with existing literature, this finding raises important equity considerations for older adults living with dementia in assisted living facilities located in rural and remote regions. ^26^ Rural assisted living facilities house fewer older adults and are more likely to have deficiencies in care provision than urban ones, including challenges with retaining appropriate care staff and resources to meet the needs of residents. ^27^ The use of videoconferencing and other information technology resources to provide dementia care should be considered to improve access to care for individuals living with dementia in rural and remote areas. ^28^

Most assisted living facilities that provided a dementia care program had capacity for more than 140 older adults and had more than 115 suites. Current practices for designing settings specifically for older adults living with dementia emphasize larger spaces that are not characteristic of institutionalized congregate care, ^29^ and the presence and statistically significant association of many suites in assisted living facilities that provide a dementia care program aligns with the literature. In addition, this may indicate that many assisted living facilities that provide a dementia care program are large complexes, likely attributed to chains.

Given the challenges that older adults living with dementia face with respect to eating, ^5^ it is expected that assistance with feeding would be a prevalent care service offered alongside a dementia care program in an assisted living facility. Moreover, the complex and intersecting care needs of older adults living with dementia, which includes polypharmacy, ^30^ underscores the need for a physician to monitor and evaluate the care plan. As such, the prevalence of medical services in assisted living facilities that offer a dementia care program is also expected.

As the assisted living sector is privately financed in Ontario, this study makes an important contribution to the literature to define the sector by modeling facility-level characteristics associated with the provision of a dementia care program. The findings are relevant to clinicians and policymakers actively considering dementia care options in communities to support older adults living with dementia and their caregivers. Family caregivers and consumers of assisted living services will also be interested in these findings to inform their decisions for care.

A limitation to this study is that the fees older adults residing in assisted living facilities pay each month and per care service could not be included in the adjusted model. This is due, in part, to the inability to capture this information in existing administrative health system data. Moreover, these data are not publicly available on the websites of assisted living facilities, through their member associations, or available to the RHRA through regulatory reporting requirements. Another limitation is that this study is descriptive; as such, no causal or temporal claims can be made about the associations between the facility-level characteristics of assisted living facilities and the provision of a dementia care program. As with all secondary analyses of data, the data used in this study are susceptible to misclassification bias.

## CONCLUSION

This study identified and compared facility-level characteristics of licensed assisted living facilities that provided a dementia care program to those that did not in Ontario, Canada in 2018. Assisted living facilities that offered a dementia care program housed more older adults and provided more care services. Future research might consider investigating the underlying differences in populations between residents of these facilities and their health outcomes attributed to care services provided in assisted living facilities.

## Data Availability

The dataset from this study is held securely in coded form at ICES. While data sharing agreements prohibit ICES from making the data set publicly available, access may be granted to those who meet pre-specified criteria for confidential access, available at www.ices.on.ca/DAS. The full data set creation plan and underlying analytic code are available from the corresponding author upon request, understanding that the programs may rely upon coding templates or macros that are unique to ICES.

## Acknowledgments

This study was supported by ICES, which is funded by an annual grant from the Ontario Ministry of Health and Long-Term Care (MOHLTC). The opinions, results and conclusions reported in this paper are those of the authors and are independent from the funding sources. No endorsement by ICES or the Ontario MOHLTC is intended or should be inferred.
We extend our gratitude to Paul Pham, Adriane Castellino, and Chloe Ma at the Retirement Homes Regulatory Authority for sharing the registry of licensed assisted living facilities. We also extend our thanks to Nathan M. Stall for his comments on an earlier draft of this manuscript.

## Appendix

**Appendix Table 1.** Detailed Descriptions of Exposures

| **Variable** | **Description** |
| --- | --- |
| Urban | Based on the PCCF flag, which is a specialized macro for use with health system administrative datasets containing postal codes. This macro is based on 2016 Census information, flags communities with a population less than 10,000 individuals as rural, and it includes related data from Canada Post Corporation. |
| Facility Capacity | Resident capacity of the assisted living facilities. This variable was transformed into quartiles for ease of interpretation. |
| Total Suites | Number of suites in the assisted living facility. This variable was transformed into quartiles for ease of interpretation. |
| Chain Status | Binary variable specifying whether the assisted living facility is part of a chain. Chains are defined as two or more assisted living facilities owned and operated by the same entity. |
| Residential Home | Binary variable specifying whether the assisted living facility is a residential home. Residential homes are not facilities and are located in residential areas. |
| Co-Located with a Nursing Home | Binary variable specifying whether the assisted living facility is co-located with a nursing home in the same building or land site. |
| Assistance with Bathing | Binary variable specifying whether the assisted living facility provides bathing respecting privacy and preferences. |
| Assistance with Hygiene | Binary variable specifying whether the assisted living facility provides grooming, oral care, foot care, and care for fingernails. |
| Assistance with Ambulation | Binary variable specifying whether the assisted living facility provides assistance with mobility devices and transferring and positioning techniques. |
| Assistance with Feeding | Binary variable specifying whether the assisted living facility provides assistance to residents to eat and drink safely, including assistance inserting and removing dentures. |
| Assistance with Dressing | Binary variable specifying whether the assisted living facility provides assistance with dressing, consistent with time of day and weather conditions. |
| Continence Care | Binary variable specifying whether the assisted living facility provides continence products and measures to prevent constipation. |
| Skin and Wound Care | Binary variable specifying whether the assisted living facility provides interventions and routine care to maintain the integrity of the resident's skin, prevent wounds and infections, and other preventive measures (e.g., physiotherapy, etc.). |
| Provision of Meals | Binary variable specifying whether the assisted living facility provides meals supporting good nutrition standards, food service workers, and individualized meals, if required. |
| Administration of Medications | Binary variable specifying whether the assisted living facility stores medications and has regulated health professionals providing medication to residents consistent with their needs and care plans. |

**Appendix Table 1.** Detailed Descriptions of Exposures
| **Variable** | **Description** |
| --- | --- |
| Pharmacist Services | Binary variable specifying whether the assisted living facility provides services from a pharmacist regulated by the Ontario College of Pharmacists. |
| Nursing Services | Binary variable specifying whether the assisted living facility provides services from a nurse regulated by the Ontario College of Nurses. |
| Medical Services | Binary variable specifying whether the assisted living facility provides services from a physician regulated by the Ontario College of Physicians and Surgeons. |

Detailed descriptions of the care services are found in O. Reg. 166/11: GENERAL under *Retirement Homes Act, 2010*, S.O. 2010, c. 11. Available at: https://www.ontario.ca/laws/regulation/110166#BK47

**Appendix Table 2.**
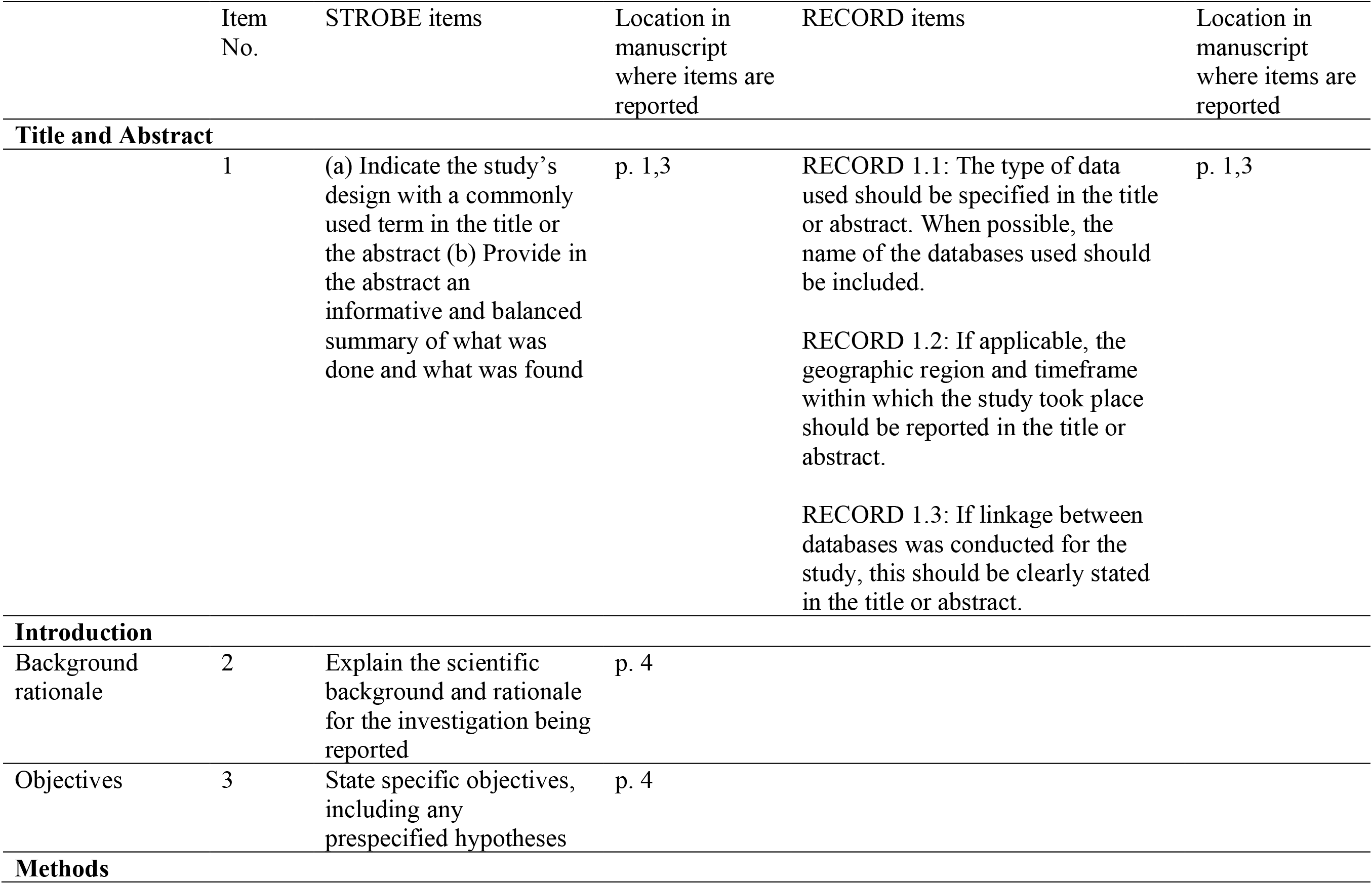
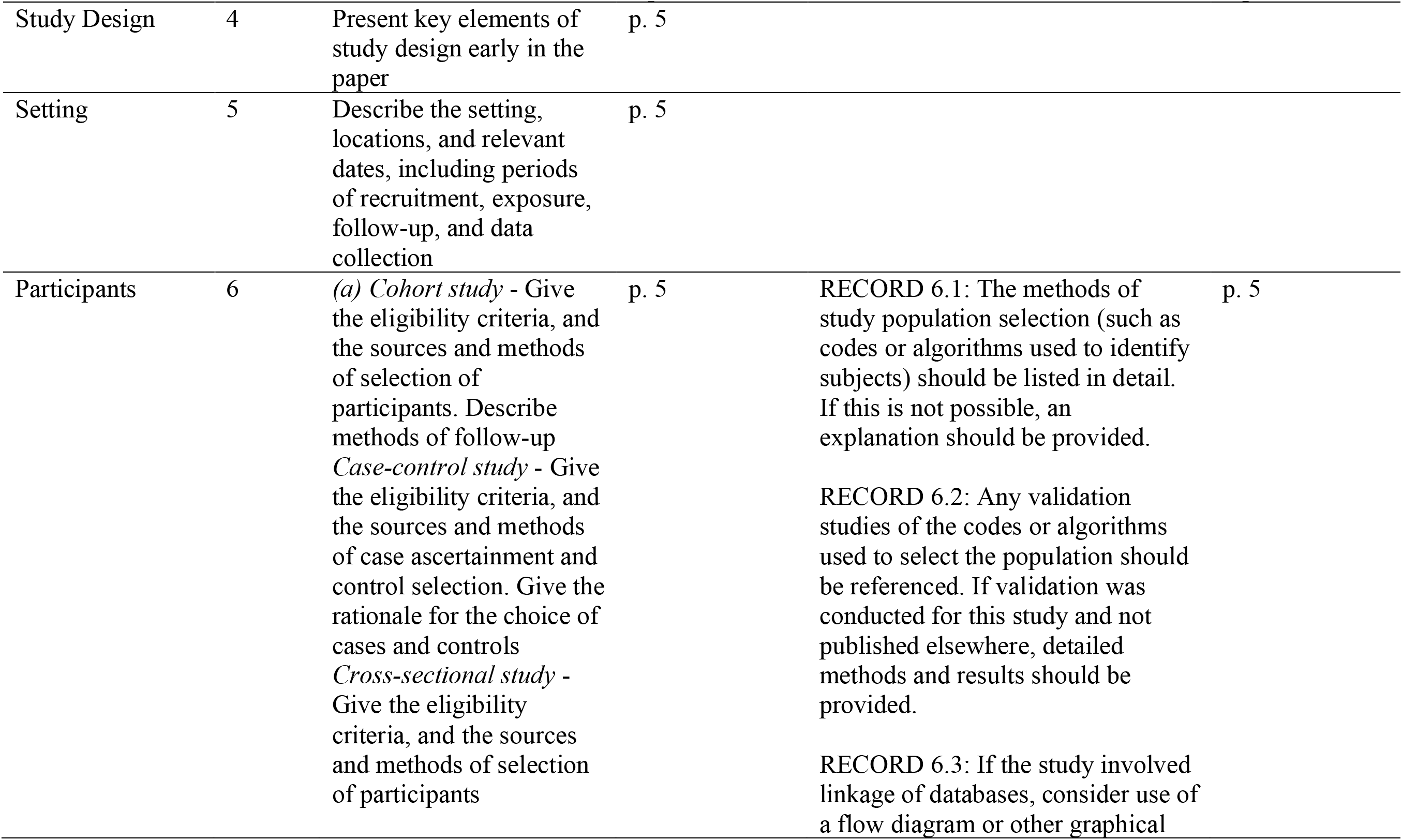
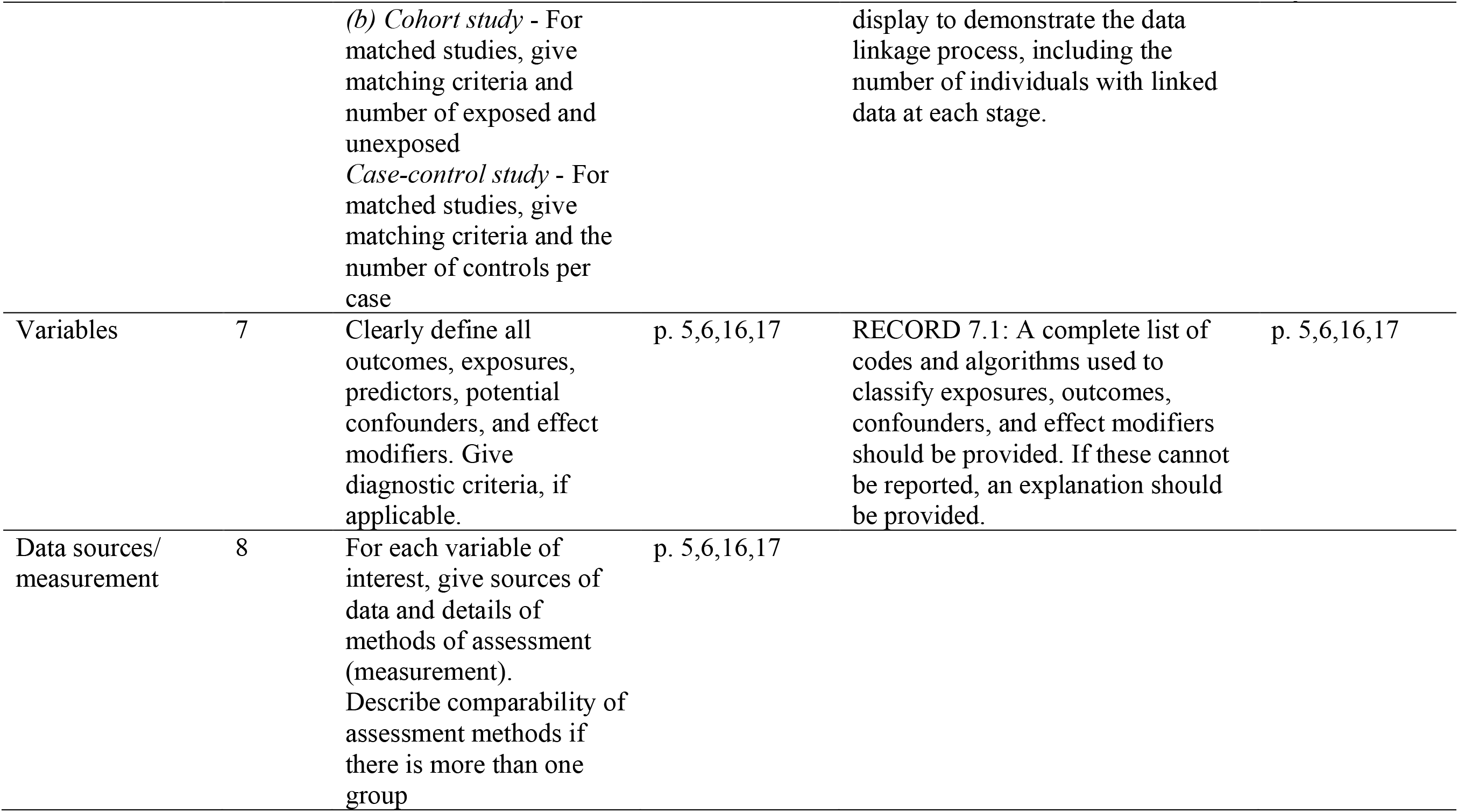
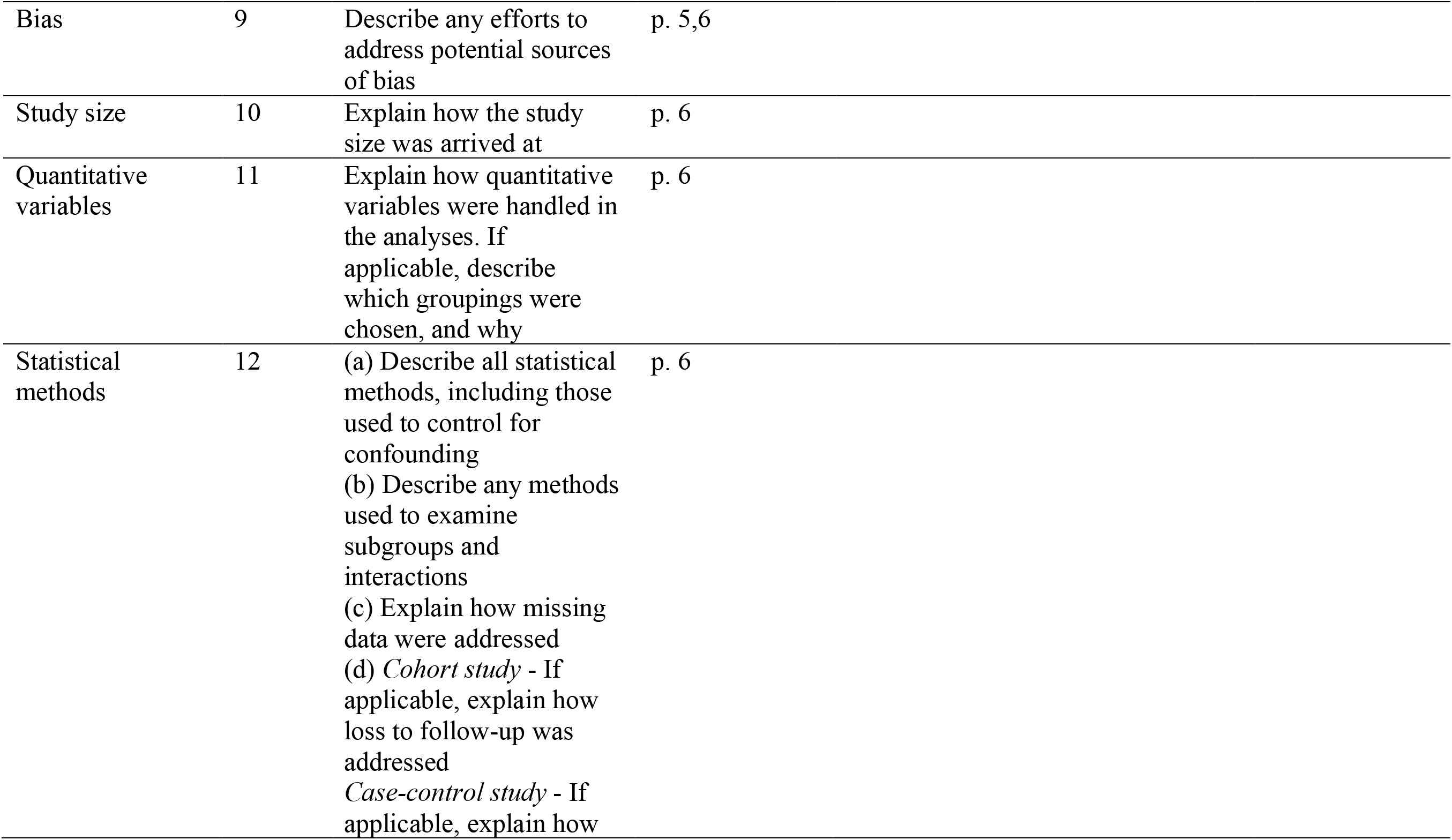
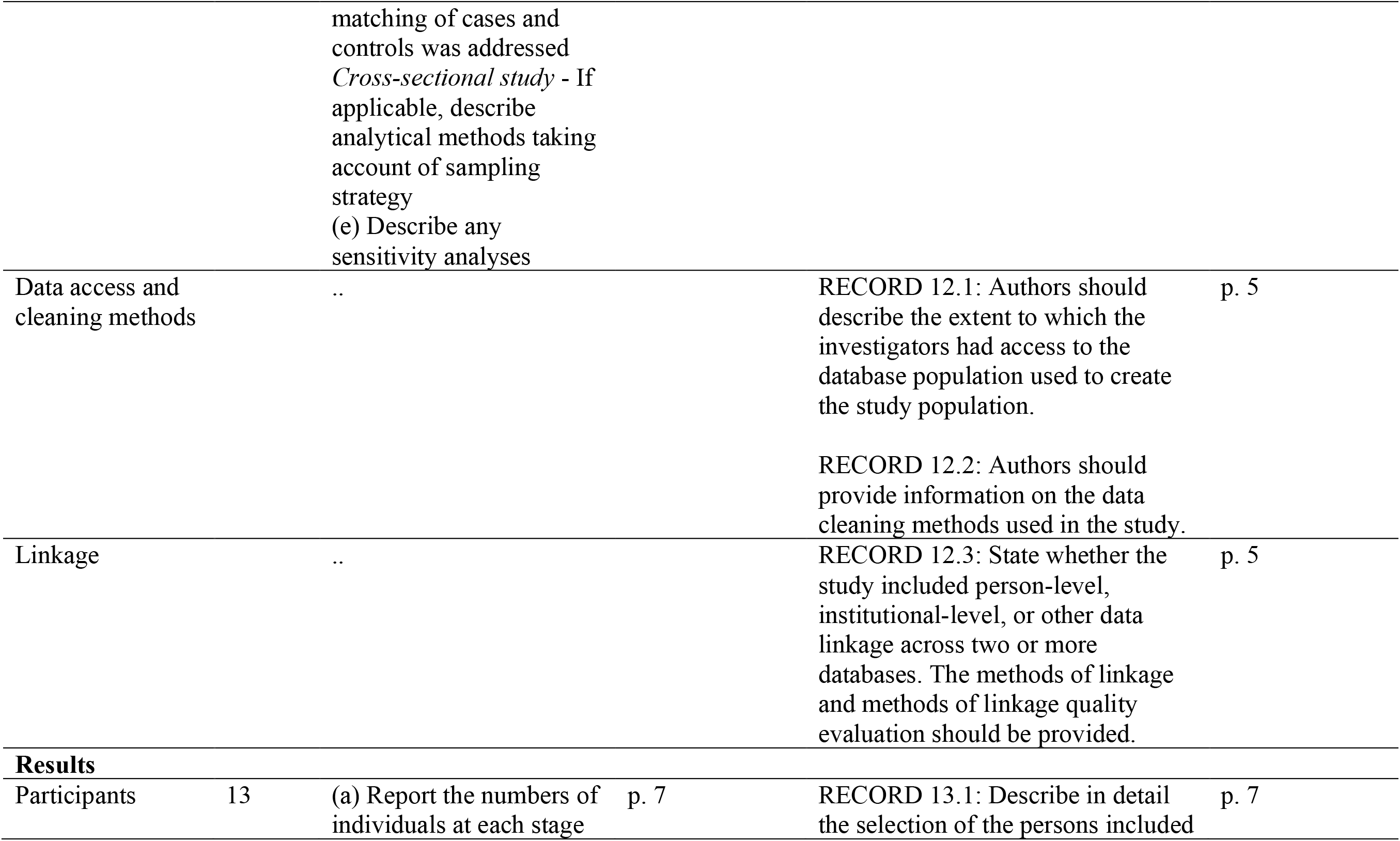
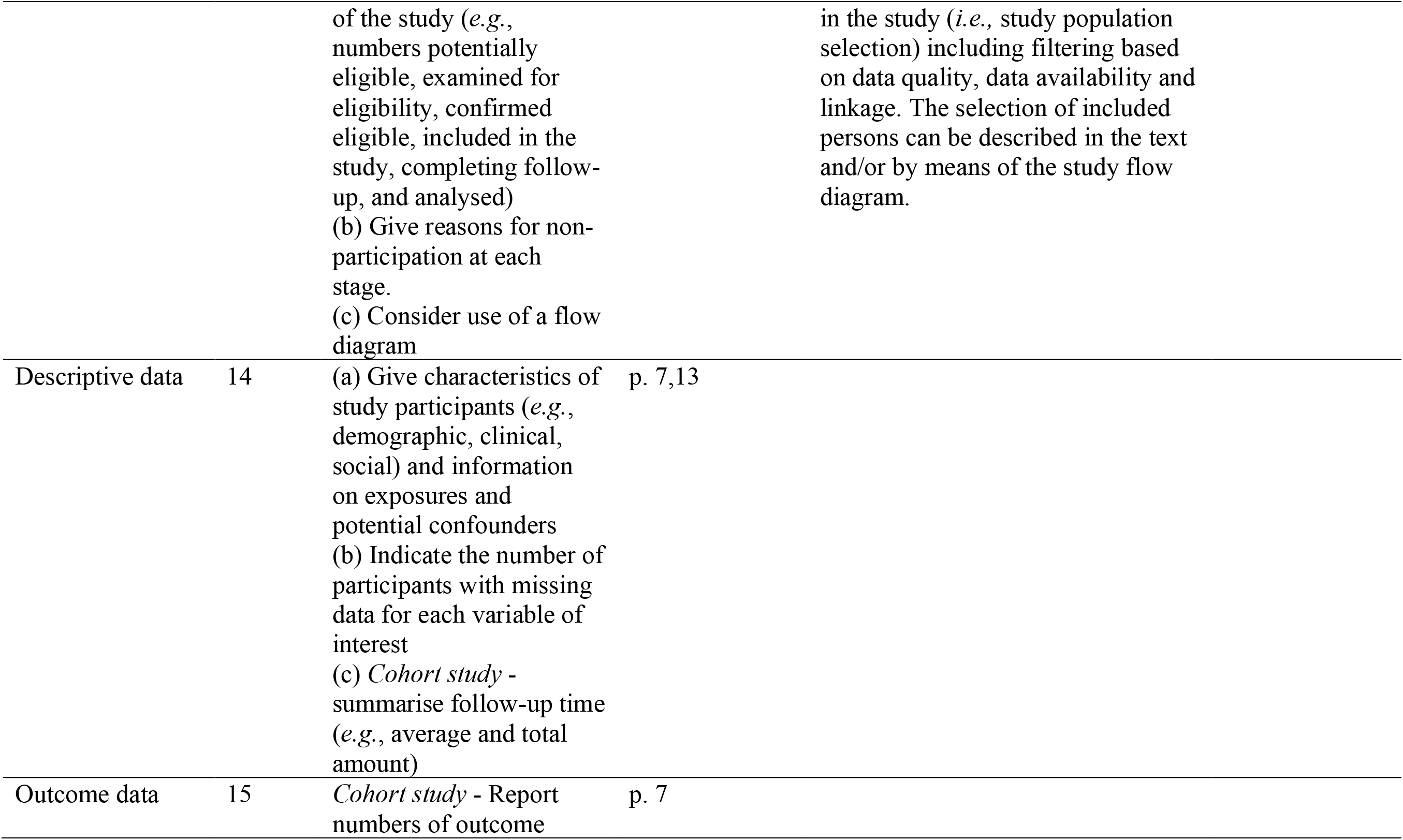
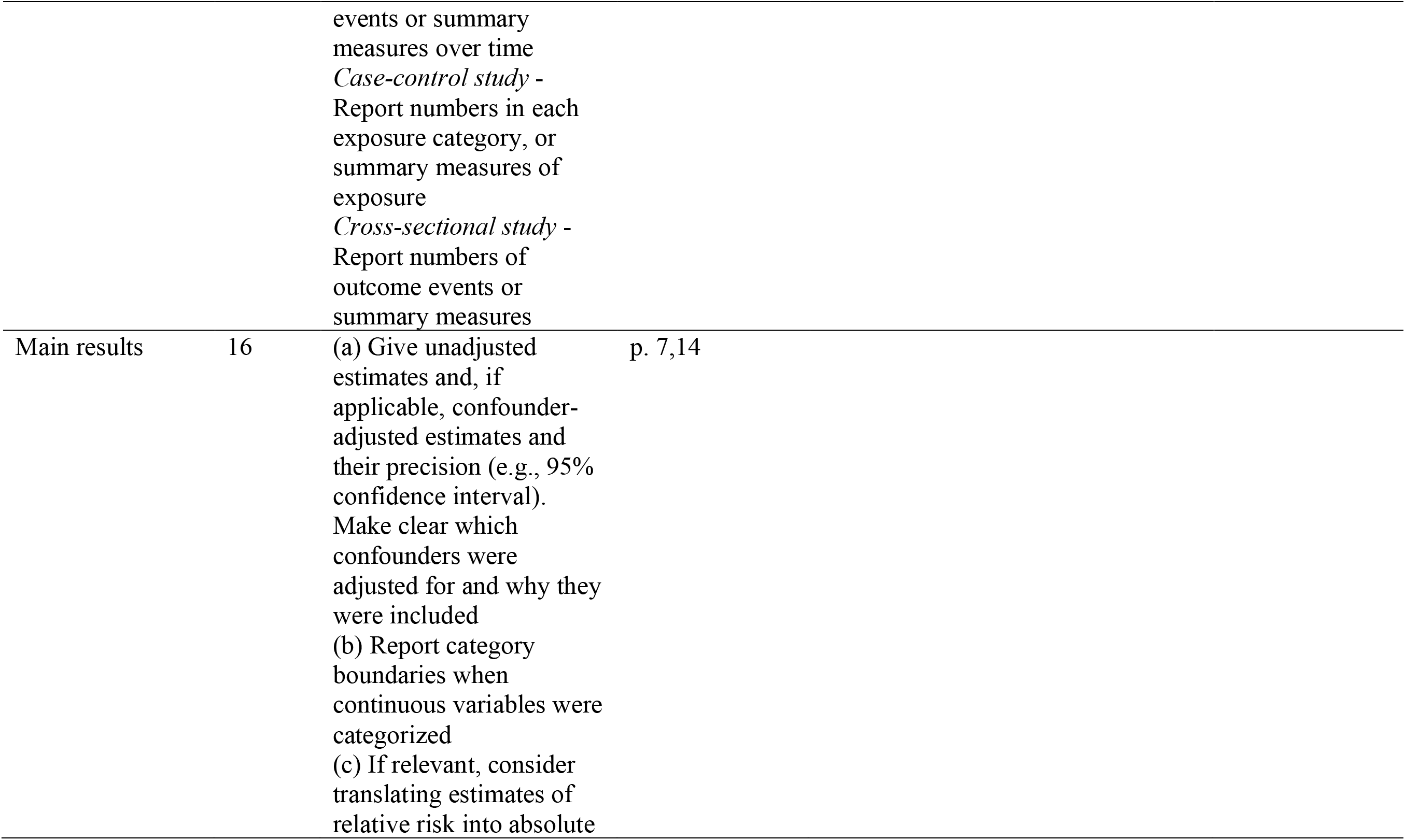
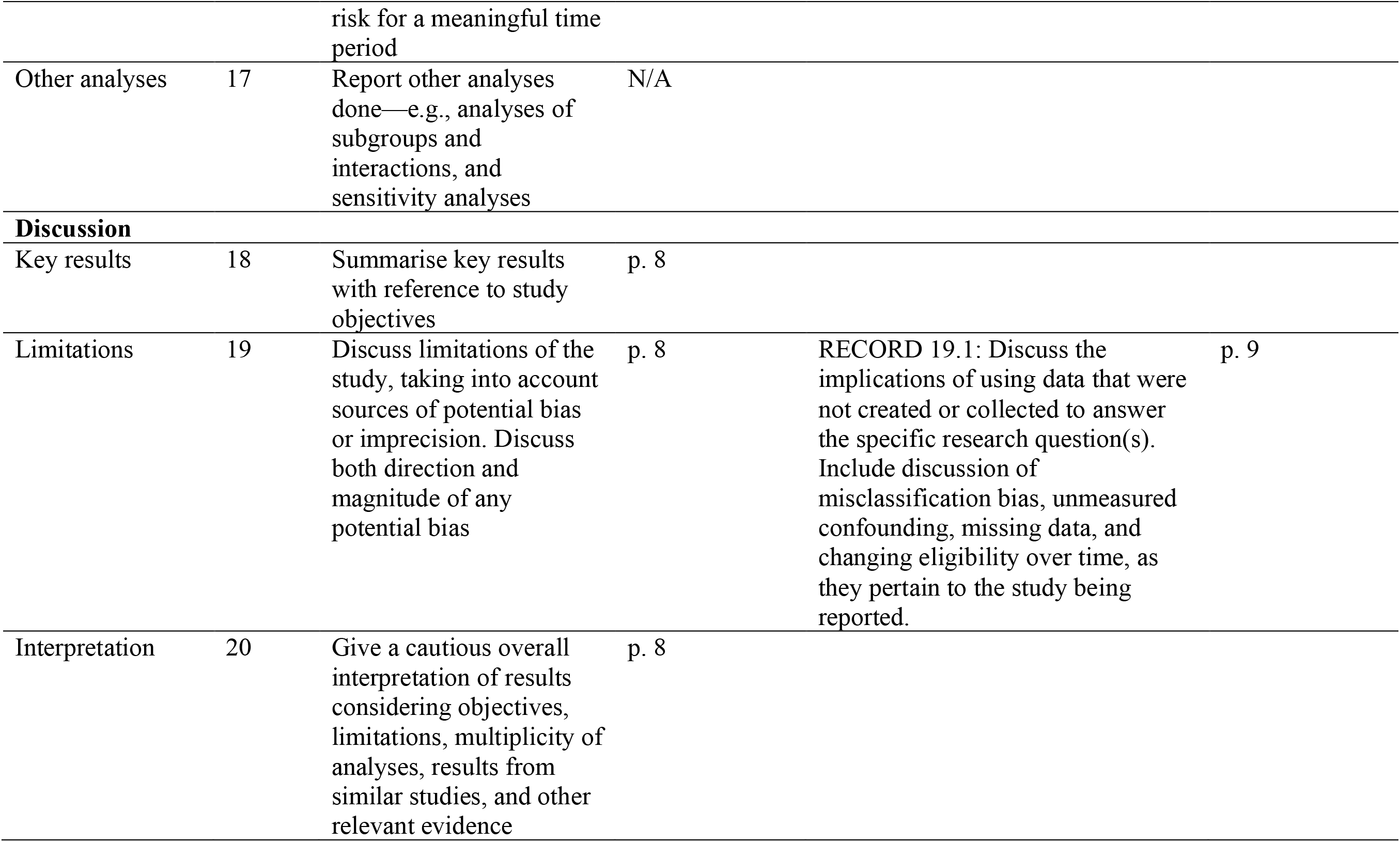
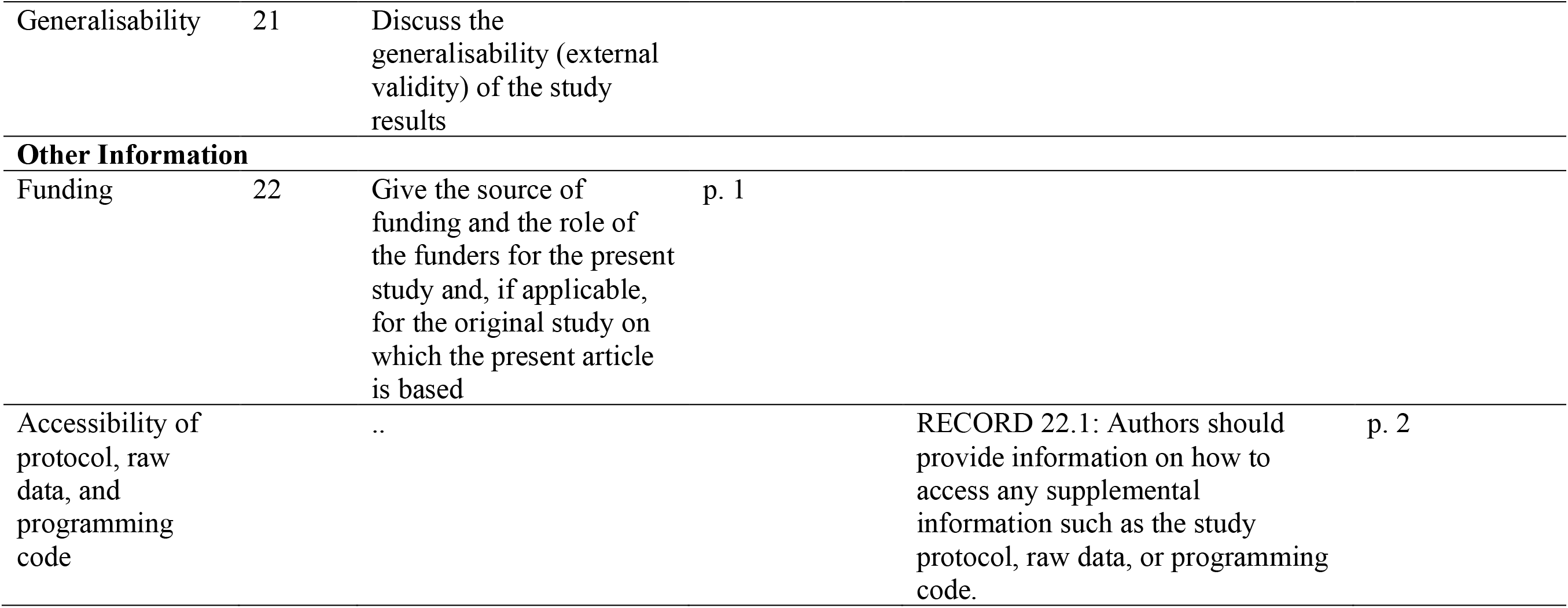
RECORD Checklist

